# Characterizing obesity-HFpEF in younger acute respiratory failure patients

**DOI:** 10.1101/2025.05.27.25327926

**Authors:** Jia Chen, Frank H Annie, Mohammed Nor, Sudha Arulalan, Dany Tager, Elie Gharib, Anand Chockalingam

## Abstract

**Background:** Hypoxic hypercapnic respiratory failure is a major cause of acute hospitalization. Cardiopulmonary insults can lead to respiratory distress and, if untreated, respiratory failure. West Virginia has the highest prevalence of obesity in the nation, accounting for the higher incidence of heart failure with preserved ejection fraction (HFpEF). This diagnosis is elusive in younger patients below age 65 years because echocardiograms are often normal and invasive hemodynamics are not widely measured. H2FPEF score, “obesity age” (BMI + age), and newly described HFpEF-ABA scores may predict obesity-related HFpEF in acute respiratory failure settings. Given the global rise in obesity, early identification of HFpEF is critical to optimizing respiratory failure care.

**Purpose:** This cohort study investigates the burden of obesity-related HFpEF in patients younger than 65 years old presenting with respiratory failure.

**Methods:** The study included patients ≤65 years of age admitted to Charleston Area Medical Center from 1/2023-12/2023 with respiratory failure, BMI > 30 kg/m2, and EF > 50%. Patients with specific cardiomyopathies or identifiable triggers for for respiratory failure were excluded. “Premature HFpEF” was defined as ‘obesity years’ >100. Demographic data on BMI, echocardiographic and hemodynamic findings were analyzed. Logistic regression was used to assess the relationship between premature HFpEF and respiratory failure, adjusting for confounders.

**Results:** Among 44 patients with respiratory failure, 64% exhibited premature obesity-related heart failure with preserved ejection fraction (HFpEF), characterized by a mean age and body mass index (BMI) of 111.5 kg/m2-years. These patients demonstrated elevated left ventricular end-diastolic pressure (LVEDP) exceeding 18 mmHg during cardiac catheterization. H2FPEF score could not be measured due to obesity limiting Doppler echocardiographic studies. The HFpEF -ABA score was > 80% and ‘obesity age’ was > 95 in 85% of the cohort.

**Conclusion:** Early recognition of obesity-related HFpEF enables healthcare providers to tailor individualized treatment regimens. Preventive strategies focused on obesity reduction can reverse associated comorbidities, improve patient outcomes, and lower healthcare costs in the long run.

## Background

Hypoxic hypercapnic respiratory failure is a major cause of rising inpatient admissions. Over the period of 2002-2017, the incidence of respiratory failure has increased 83% from 249 to 455 cases per 100,000 adults (1). This can lead to mechanical ventilation, resulting in longer hospitalizations averaging 10.5 days and costing over $150,000.

West Viriginia has the highest prevalence of obesity in the nation at 37.7% and this has largely impacted the rates of respiratory failure (2). Obesity has been associated with reduced airflow leading to airway hypersensitivity and increased risk for the development of pulmonary diseases such as obstructive sleep apnea, obesity hypoventilation syndrome and pulmonary hypertension (3). These physiological changes of obesity burden the lungs contributing to hypoxic and/or hypercapnic failure.

Longitudinal population cohorts have clearly demonstrated the increasing incidence of heart failure proportional to higher body mass index (BMI) (4). Excess adipose tissue in obesity increases body surface area and therefore raises metabolic demand, blood volume and cardiac output (5). The sequalae of increased cardiac output raises left ventricular wall stress, leading to left ventricular hypertrophy. Therefore, having to meet the increased demands of the body strains the ventricle, thus impairing relaxation leading to diastolic dysfunction (6). Younger patients with higher obesity burden are at risk of developing obesity related HFpEF prematurely with resultant complications.

Respiratory distress from cardiopulmonary insults has burdened our healthcare system and contributed to substantial health care costs. Given the global rise in obesity, early identification of HFpEF is critical to optimizing respiratory failure care. HFpEF is typically diagnosed in older patients. In younger patients below 65 years of age, obesity alone may be sufficient risk predictor for HFpEF (7). Typical echocardiographic features of left atrial dilation and Doppler diastolic dysfunction and BNP elevation are often absent in premature obesity related HFpEF settings. We proposed “obesity years” (age + BMI > 100) to gauge the risk for obesity-related HFpEF in patients younger than 65 years of age. In many instances, cardiac catheterization with fluid challenge to elicit LVEDP increase to over 18mmHg is required to definitively diagnose HFpEF. Mayo described H2FPEF score in 2018 and recognizing the challenges with obtaining echo variables, validated HFpEF-ABA score in 2024. We analyze the clinical characteristics and predictive utility of these scoring systems in respiratory failure patients with obesity-HFpEF.

## Methods

This retrospective cohort study was conducted at Charleston Area Medical Center and included adult patients aged 18 to 65 years who were admitted between January 1, 2023, and December 31, 2023, with a diagnosis of acute respiratory failure. Eligible patients were identified using the institution’s electronic health record (EHR) system. Inclusion criteria were age ≤65 years, a primary diagnosis of hypoxemic (PaO2 <60 mmHg) or hypercapnic (PaCO2 >45 mmHg) respiratory failure requiring supplemental oxygen or ventilatory support, body mass index (BMI) greater than 30 kg/m^2^, and preserved left ventricular ejection fraction (LVEF >50%) on echocardiography obtained during hospitalization. Patients were excluded if they had a history of non-obesity-related cardiomyopathies, acute myocardial infarction or active myocarditis during admission, primary pulmonary causes of respiratory failure with an identifiable trigger (such as sepsis, pneumonia, or massive pulmonary embolism), prior diagnosis of heart failure with reduced ejection fraction, or significant valvular or congenital heart disease.

Demographic and clinical data were abstracted from the EHR, including age, sex, race, BMI, comorbidities, and relevant laboratory values. Echocardiographic data were reviewed for left ventricular wall thickness, left atrial size, diastolic function parameters, and ejection fraction. When available, invasive hemodynamic measurements from cardiac catheterization, including left ventricular end-diastolic pressure (LVEDP), were collected. Obesity-related HFpEF was defined as an “obesity years” score (age plus BMI) greater than 100. The H2FPEF and HFpEF-ABA scores were calculated for each patient when data permitted, following previously published methods; inability to obtain Doppler measures due to obesity was recorded.

The primary outcome was the prevalence of premature, obesity-related HFpEF among patients under 65 years of age admitted with acute respiratory failure. Secondary outcomes included the proportion of patients with left ventricular hypertrophy, left atrial enlargement, elevated LVEDP (>18 mmHg), and high-risk HFpEF-ABA scores (>80%). Logistic regression analysis was performed to assess the association between premature HFpEF (obesity years >100) and respiratory failure, adjusting for confounders including age, sex, and comorbidities. Continuous variables were reported as means with standard deviation or medians with interquartile range, and categorical variables as counts and percentages. Statistical significance was defined as a two-sided p-value less than 0.05. Data analysis was performed using Stata 16. The study was approved by the Charleston Area Medical Center Institutional Review Board, with a waiver of informed consent due to the retrospective nature of the analysis.

## Results

From January 2023 to December 2023, we identified 44 acute respiratory failure hospitalized patients with severe obesity with normal systolic function (figure1). Twenty-five of these 44 patients had left ventricular hypertrophy and left atrial enlargement. These 44 patients had a variety of hemodynamic dysfunctions: 17 patients were found to have pulmonary hypertension, 28 patients had elevated left ventricular end-diastolic pressure (LVEDP), and 20 patients had high cardiac output. As fluid challenge is not routinely performed at our institution, these patients demonstrated elevated LVEDP exceeding 18 mmHg during cardiac catheterization. H2FPEF score was 99 and 98% probability for HFpEF in 2 of the patients – but could not be calculated in the remainder due to obesity limiting Doppler echocardiographic studies. The HFpEF -ABA score was > 80% and ‘obesity age’ was > 95 in 85% of the cohort.

**Figure1.**
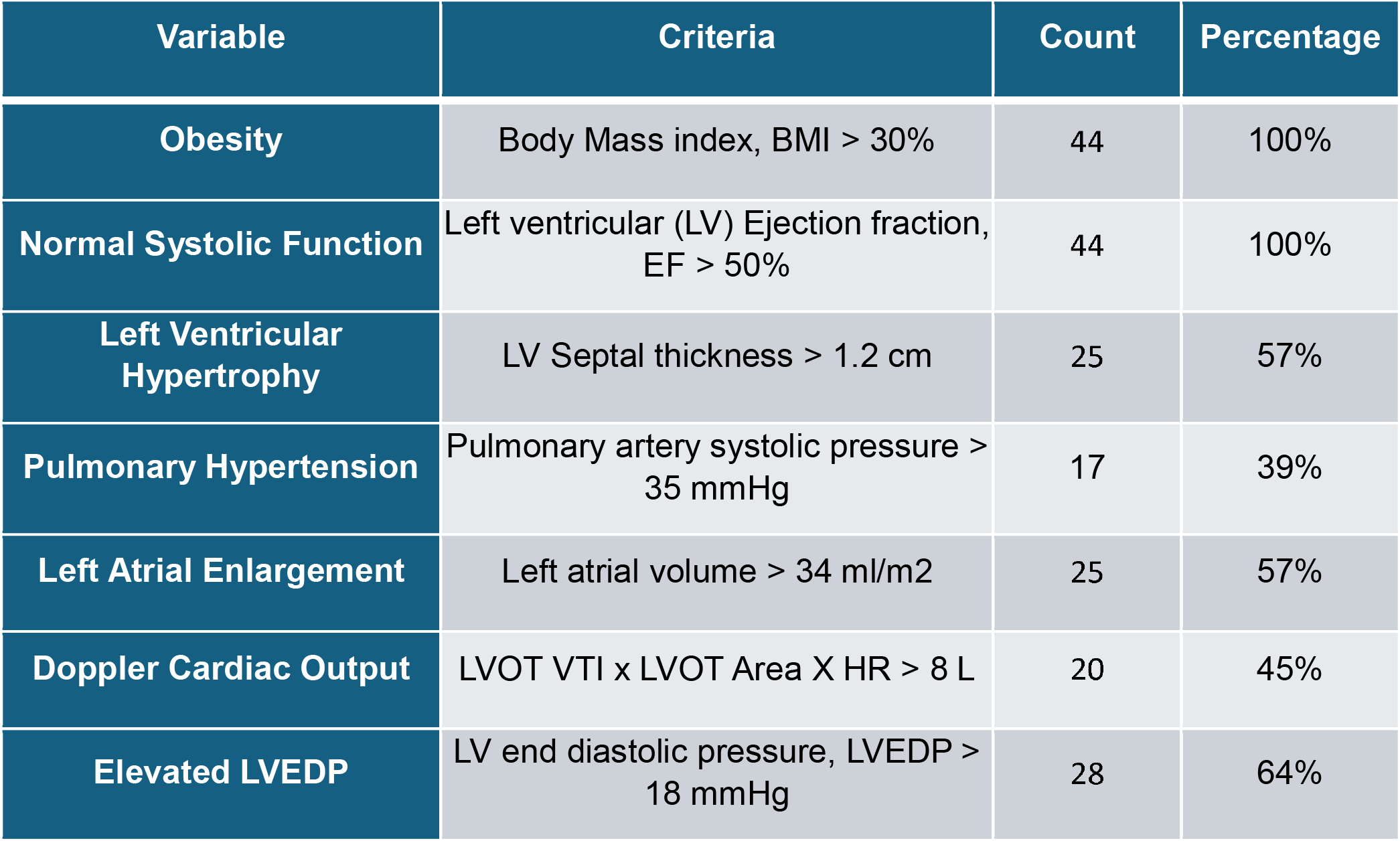
Prevalence of clinical predictors of heart failure with preserved ejection fraction (HFpEF) among consecutive young patients < 65 years age hospitalized with acute respiratory failure.

## Discussion

This study in a multidisciplinary referral hospital shows there is a significant burden of obesity related heart failure among younger patients with acute respiratory failure. Non-invasive diastolic function estimation tends to be within normal range oftentimes as does BNP in younger obese individuals. Even when cardiologists get involved, several challenges make this diagnosis elusive.

We are increasingly recognizing obesity as a significant risk factor for HFpEF, with studies indicating that up to 85% of HFpEF patients are overweight or obese (8). Obesity has long been associated with reduced airflow leading to hypoxia and hypercarbic respiratory failure through several mechanisms. The excessive adipose tissue found in the chest wall and abdomen limits breathing by causing impaired gas exchange, leading to hypoxia. In addition, obesity also reduces lung volume, especially residual capacity and expiratory reserve volume, which leads to rapid desaturation. The combination of increased work of breathing caused by excessive weight on the chest wall, combined with the response to hypercapnia and hypoxia, exacerbates respiratory failure. (9)(10)

The growing concerns of having higher body mass index (BMI) therefore raises metabolic demand, blood volume and cardiac output (5). Excess adipose tissue in obesity alters the cardiac structures and causes HFpEF. This development of HFpEF involves several key factors, including concentric left ventricular hypertrophy, increased left ventricular pressures, and left atrial dilation are the sequala of elevated left ventricular (LV) pressure to accommodate the increased volume in obesity. Long term elevated LV pressure causes pressure to build up in the left atrium and ultimately to atrial dilation and elevated pulmonary pressures. As the chronicity of the elevated left sided pressure continues, the pulmonary vasculature adapts by increasing resistance, leading to the development of pulmonary hypertension (11).

We believe obesity-HFpEF leading to respiratory failure involves complex interactions between obesity, cardiac function, and respiratory mechanisms. Obesity causes altered pulmonary mechanics by reducing lung volume, residual capacity, and expiratory reserve volume. The reduction of residual capacity and expiratory reserve volume causes expiratory flow limitations and generates intrinsic positive end-expiratory pressure. This increased work of breathing is further exacerbated by the increased load of thoracoabdominal pressures. However, such patients also have cardiac remodeling exhibiting increased left ventricular muscle mass with LV hypertrophy, which increases pressures in the LV. These changes create longstanding resistance in the pulmonary vasculature, leading to pulmonary hypertension. (12)

In our 44 subjects, 25 have elevated LVEDP confirming obesity-HFpEF contribution to the clinical picture of shortness of breath and respiratory distress. But diagnosis in such population is a challenge in terms of biomarker screening, noninvasive imaging as well as right and left heart catheterization. It requires an invasive pressure reading of the left ventricle to diagnose the condition before unloading with diuretics ideally. We routinely re-evaluate for LVEDP increase over 18mmHg with 2 bolus infusions of 500cc normal saline rapidly to elicit occult cases. Obesity phenotype HFpEF is under-recognized, and clinicians should consider catheterization to confirm LV pressure if patients have multiple comorbidities associated with obesity. This highlights importance of early recognition and a multidisciplinary team approach for such patient populations in optimization of cardio-respiratory care and tailored treatment.

## Limitation

Data are gathered from a single center with a limited number of patients and a lack of randomization to control confounding variables. Given the increasing prevalence of obesity, particularly in West Virginia, it is essential to conduct rigorous studies that encompass a larger population. Such studies will enable clinicians to adopt multidisciplinary approaches and mitigate the burden of obesity, which significantly impacts a substantial proportion of our patient population.

## Data Availability

All data produced in the present work are contained in the manuscript

## References

1. Kempker JA, Abril MK, Chen Y, Kramer MR, Waller LA, Martin GS. The epidemiology of respiratory failure in the United States 2002–2017: a serial cross-sectional study. Crit Care Explor. 2020;2(6):e0128. https://pubmed.ncbi.nlm.nih.gov/32695994/

2. West Virginia Department of Health and Human Resources (WV DHHR). Weight status and chronic disease statistics. Fast Facts. https://dhhr.wv.gov/hpcd/data_reports/pages/fast-facts.aspx (Use this link for official state statistics; the anchor may not always work, but the page is correct.)

3. Costello RW. Obesity and respiratory disease. Breathe (Sheff). 2023;19(1):220263. https://breathe.ersjournals.com/content/19/1/220263.short

4. Yusuf S, Hawken S, Ounpuu S, et al. Effect of potentially modifiable risk factors associated with myocardial infarction in 52 countries (the INTERHEART study): case-control study. Lancet. 2004;364(9438):937–52. https://www.nejm.org/doi/full/10.1056/NEJMoa020245

5. Carbone S, Lavie CJ, Arena R. Obesity and heart failure: focus on the obesity paradox. Heart Fail Clin. 2013;9(3):269–277. https://www.sciencedirect.com/science/article/abs/pii/S1871403X1300224X

6. Lavie CJ, Osman AF, Milani RV, Mehra MR. Body composition and prognosis in chronic systolic heart failure: the obesity paradox. JACC Heart Fail. 2013;1(2):93–102. https://www.jacc.org/doi/full/10.1016/j.jchf.2013.01.006

7. Norton GR, Woodiwiss AJ. The obesity paradox and heart failure with preserved ejection fraction: A matter of fat distribution? Front Cardiovasc Med. 2022;9:821829. https://www.frontiersin.org/articles/10.3389/fcvm.2022.821829/full

8. Anker SD, Butler J, Filippatos G, et al. Empagliflozin in heart failure with a preserved ejection fraction. N Engl J Med. 2021;385(16):1451–1461. https://www.nejm.org/doi/full/10.1056/NEJMoa2107038

9. Zhang Y, Zhang J, Butler J, et al. Contemporary epidemiology, management, and outcomes of patients hospitalized for heart failure in China: results from the China Heart Failure Registry Study. Front Cardiovasc Med. 2023;10:10292783. https://www.ncbi.nlm.nih.gov/pmc/articles/PMC10292783/

10. Sanchis-Gomar F, Perez-Quilis C, Lavie CJ, et al. Obesity and heart failure with preserved ejection fraction: an update. Eur Respir Rev. 2019;28(151):180097. https://err.ersjournals.com/content/28/151/180097

11. Chockalingam A. “Obesity-years” burden may predict reversibility in heart failure with preserved ejection fraction. Front Cardiovasc Med. 2022;9:830097. https://www.frontiersin.org/articles/10.3389/fcvm.2022.830097/full

12. Obokata M, Reddy YNV, Pislaru SV, Melenovsky V, Borlaug BA. Evidence supporting the existence of a distinct obese phenotype of heart failure with preserved ejection fraction. Circulation. 2017;136(1):6–19. https://pubmed.ncbi.nlm.nih.gov/28381470/

